# Integrated epidemiology and toxicology reveals the protective effects of TMAO against chemical neurotoxicity in children

**DOI:** 10.64898/2026.07.02.26357012

**Authors:** V.C. de Leeuw, L. Maitre, C.T.M. van Oostrom, E. Renard-Dausset, A. Anguita, L. Chatzi, M. Coen, R. Gražulevičienė, B. Heude, J. Ibarluzea, J. Julvez, H.C. Keun, A.H. Piersma, L. Santa-Marína, S. Marquez, M. Ruiz-Rivera, M. Subiza-Perez, A.L. Brantsæter, M.B. Toledano, M. Vrijheid, J. Wright, E.V.S. Hessel, L. Hoyles, S. McArthur

## Abstract

Interest in microbiota–host co-metabolism and the effects of its derived co-metabolites on biological processes is increasing rapidly. In addition to their demonstrated associations with mammalian metabolic health and cognition, microbiota–host co-metabolites (MHCMs) represent lifelong contributors to the endogenous exposome. We have previously shown the MHCM trimethylamine *N*-oxide (TMAO) to exert beneficial effects on murine blood–brain barrier integrity and cognition. Here we investigated whether these positive neural effects of TMAO extended to humans, analysing how TMAO exposure associates with neurodevelopmental outcomes in children and whether an *in vitro* human neuronal–astrocyte co-culture could contribute to further investigation of the underlying mechanism(s) and neuronal processes related to these associations. In a cohort study of childhood mental health (*N*=1,203), TMAO was associated with fewer internalising problems, while its precursor microbial metabolite trimethylamine was associated with more behavioural problems in both the cross-sectional and an independent longitudinal study from 1 to 15 years of age (N=630-820). Given prior associations between TMAO exposure and exposure to the environmental pollutants mercury and arsenic, we investigated how the effects of TMAO interacted with these known neurotoxicants. TMAO had a protective effect, modifying the relationship between arsenic exposure and poorer neurodevelopmental outcomes. Furthermore, TMAO activated synaptogenesis-related gene expression and was functionally protective against the negative effects of mercury in our *in vitro* model. Together, our findings emphasise the importance of interdisciplinary approaches to evaluate associations and potential pathways of MHCMs (endogenous) and environmental (exogenous) metabolites on neurodevelopment in exposome studies.

## Introduction

The human gut microbiota comprises the entirety of microorganisms that inhabit the gastrointestinal tract. Represented by ∼10^11^ microbial cells, these microbes are predicted to encode for 150 times more genes than the human genome^1^, greatly enhancing the number of potential bio-transformations that can occur within the gastrointestinal tract. This biosynthetic capacity underlies the production of a wide range of microbiota-associated metabolites that interact with human cells and biological processes throughout the body^1^ and across the lifecourse. Roles for such metabolites have been and are being identified in numerous aspects of systemic health, particularly in cardiometabolic disease and associated co-morbidities, but they are increasingly also identified as major players in the gut–brain axis, the bidirectional communication network between the gastrointestinal tract and the central nervous system^2^.

A range of structurally diverse microbiota–host co-metabolites (MHCMs) – including short-chain fatty acids^3,4^, *p*-cresol sulfate^5^, 3-phenyllactic acid^6^ and trimethylamine *N*-oxide (TMAO)^7^ – have been shown by ourselves and others to influence brain health and the response to (neuro)inflammatory stimuli. Short-chain fatty acids, for example, can regulate immune and neuroendocrine signalling and act as histone deacetylase inhibitors, providing a plausible link between microbial metabolism and transcriptional regulation in the host^8^. Tryptophan– and phenylalanine-derived microbial metabolites may influence neurotransmitter-related pathways, aryl hydrocarbon receptor signalling, oxidative stress, and neuroimmune responses^9^. TMAO, generated by host hepatic oxidation of the microbial product trimethylamine (TMA), has been implicated in oxidative stress, inflammatory signalling, endothelial function and blood–brain barrier-related processes^10^. Although mechanistic understanding of how these endogenous contributors to the exposome act is improving, to date the majority of studies of MHCMs and brain health have focused on adulthood; far less is understood about whether and how MHCMs affect child neurodevelopment, or whether they modify the developmental neurotoxicity of environmental pollutants.

Perhaps the area in which associations between the gut microbiota and neurodevelopment have been most extensively explored is autism spectrum disorder (ASD), partly because gastrointestinal symptoms are common in individuals with ASD^3^. Numerous studies have indicated that the population structure of the gut microbiota of children diagnosed with ASD differs from that of neurotypical individuals^4–6,11^, although the species variations reported are generally poorly reproducible^7^, and microbiota compositional differences are known to be heavily confounded by the distinct dietary preferences common to individuals with ASD^12,13^. Nonetheless, strategies directly targeting the gut microbiota have shown behavioural benefits in preclinical models of ASD^14^ and in a small study the non-absorbable antibiotic vancomycin temporarily reduced ADHD-related behavioural symptoms^15^, supporting the possible involvement of the gut microbiota. Since microbiota composition alone may provide limited mechanistic insight, a metabolite-focused approach may help bridge this gap by identifying microbial biochemical products that are measurable in human biospecimens, biologically active in experimental systems, and potentially linked to neurodevelopmental phenotypes.

ASD represents only one facet of childhood neurodevelopment; behavioural and cognitive outcomes encompass a much wider range of phenotypes, including attention, working memory, fluid intelligence, and internalising and externalising behaviours. Whether these are similarly influenced by MHCMs is unclear, nor is it known whether MHCMs interact with other elements of the exposome. This is particularly relevant because microbial metabolites and environmental pollutants may share dietary sources, metabolic pathways or biological targets. In our recent work in the INMA and HELIX mother-child cohorts, urinary levels of the MHCM TMAO were strongly associated with the environmental pollutants arsenic (As) and mercury (Hg) in children^16^ and pregnant women^17^. While we identified this relationship as partially driven by fish intake^18^, this did not fully explain the associations, highlighting the need to analyse more complex interactions in the context of exposome research.

Here, we investigated whether MHCMs are associated with childhood neurodevelopment and whether they interact with environmental pollutants using an integrated epidemiological–toxicological design, described in Figure 1. We combined urinary metabolomics and repeated neurodevelopmental assessments from the well-characterised HELIX and INMA birth cohorts with a human stem cell-based model of neuronal and astrocytic development. Guided by the cohort findings, we focused on TMAO and its microbial precursor TMA, and examined whether TMAO modified the effects of methylmercury and arsenic on neuronal activity *in vitro* and in cohort-based interaction analyses. This interdisciplinary approach allowed us to connect early-life exposome findings with mechanistic toxicology and to evaluate microbial co-metabolism as a potential modifier of neurodevelopmental responses to environmental pollutants.

**Figure 1.**
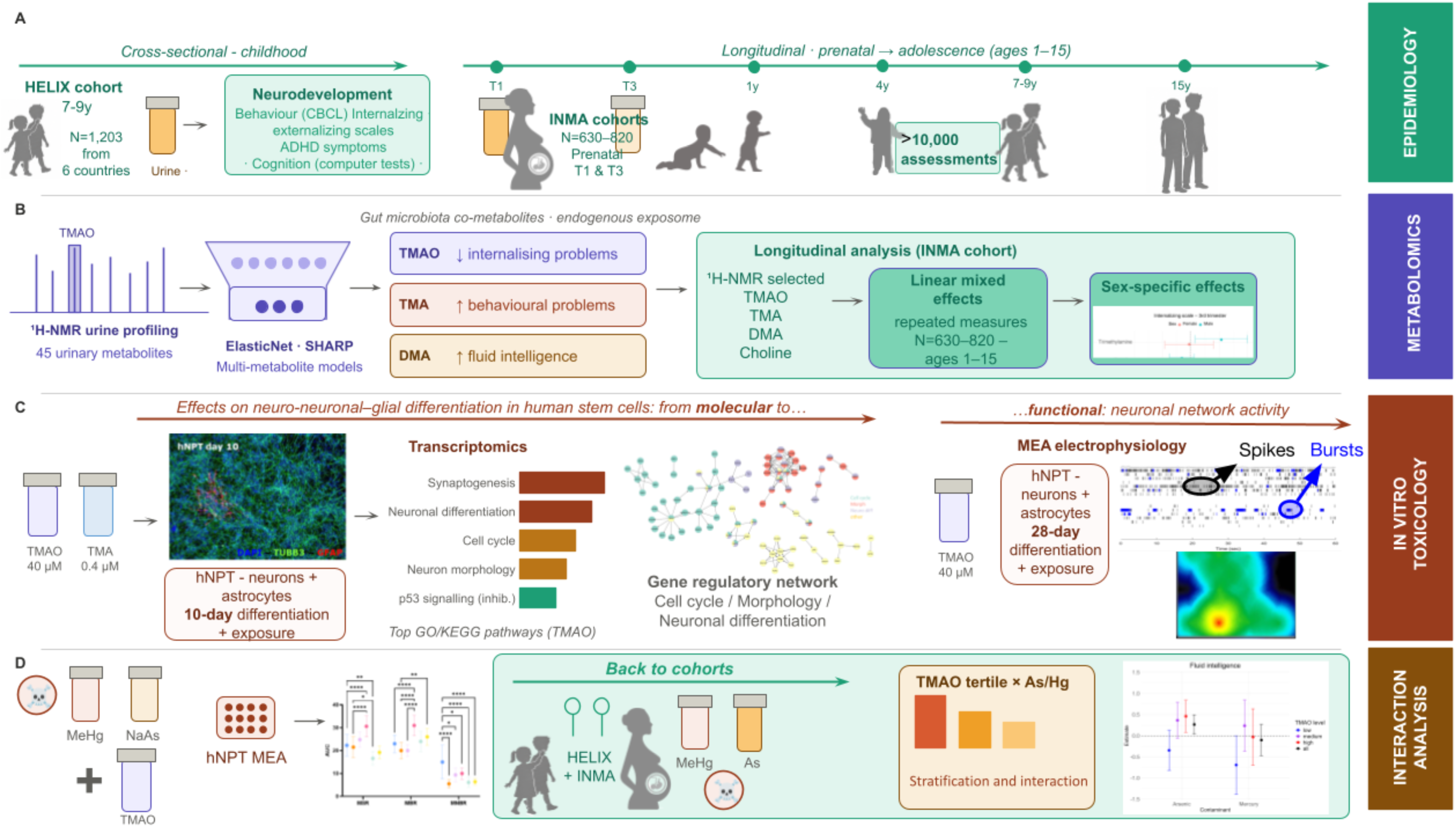
Study overview: integrated epidemiological and in vitro toxicological investigation of methylamine co-metabolites and child neurodevelopment. (A) Epidemiology. Cross-sectional associations between urinary metabolites and neurodevelopmental outcomes were assessed in children aged 6–11 years from the HELIX cohort (N=1,203, six European countries). Behavioural outcomes included internalising and externalising problems (CBCL), ADHD symptoms (Conners’ Rating Scale) and cognitive function. Longitudinal associations were assessed in the INMA birth cohorts (N=630–820, Spain), with prenatal urine collected at T1 and T3 and child outcomes assessed repeatedly from age 1 to 15 years (>10,000 assessments). **(B) Metabolomics.** Untargeted urinary ¹H-NMR metabolic profiling was performed in HELIX children (45 metabolites semi-quantified) and multi-metabolite ElasticNet models within the SHARP framework used to identify metabolites associated with neurodevelopmental outcomes. TMAO, TMA and DMA were semi-quantified by ¹H-NMR in INMA prenatal urine and tested for association with child outcomes using linear mixed-effects models, adjusting for sex, maternal education, creatinine and fish intake. All cohort analyses were sex-stratified. **(C) In vitro toxicology.** Effects of methylamines were investigated in a human neural progenitor test (hNPT) model of stem-cell-derived neurons and astrocytes. Molecular effects of TMAO (40 µM) and TMA (0.4 µM) were assessed by RNAseq over 10 days (DEGs at adjusted p-value<0.05; GO and KEGG pathway enrichment). Functional neuronal network activity due to TMAO exposure was assessed by multielectrode array (MEA) electrophysiology over 28 days. **(D) Back to cohorts: interaction analysis.** The ability of TMAO to modify neurotoxicant effects was tested in two complementary systems. In the hNPT, neurons were co-exposed to TMAO with methylmercury (MeHg) or sodium arsenite (NaAs) at environmentally relevant concentrations. In the HELIX cohort, TMAO tertiles were tested for interaction with urinary arsenic and blood mercury on fluid intelligence using likelihood ratio tests. TMAO, trimethylamine N-oxide; TMA, trimethylamine; DMA, dimethylamine; hNPT, human neural progenitor test; MEA, multielectrode array; MSR, mean spike rate; MBR, mean burst rate; MNBR, mean network burst rate; NMR, nuclear magnetic resonance; CBCL, Child Behaviour Checklist; ADHD, attention deficit hyperactivity disorder; MeHg, methylmercury; NaAs, sodium arsenite; T1/T3, first/third trimester; HELIX, Human Early-Life Exposome; INMA, INfancia y Medio Ambiente; SHARP, Stability-enHanced Approaches using Resampling Procedures

## Results

### Diet and microbiota-related metabolites are associated with child behavioural problems and cognition

We first assessed whether metabolites measured in childhood in the HELIX cohort (N=1,203) were associated with cognitive outcomes: attention, fluid intelligence (ability to solve novel reasoning problems) and working memory, adjusting for maternal education, child’s age at assessment and cohort. We also assessed behavioural outcomes: externalising behaviours (i.e. aggression/impulsivity), internalising behaviours (i.e. anxiety/low mood), total behavioural problems, and attention-deficit/hyperactivity disorder (ADHD).

Of the 45 metabolites semi-quantified in children, 21 were selected by ElasticNet regression as predictive of at least one behavioural outcome (Figure 2A). ElasticNet was used to perform variable selection while accounting for correlation among metabolites. In outcome-specific multi-metabolite models including only the selected metabolites, several nominal associations were observed with behavioural problem scores (unadjusted P < 0.05). TMAO was associated with lower internalising problems (P = 0.008), whereas its microbial precursor TMA was associated with higher internalising problems (P = 0.016). Among other statistically significantly associated metabolites, proline betaine was notable because it showed negative associations across several behavioural outcomes, including ADHD symptoms (P = 0.001), total problems (P = 0.007) and externalising symptoms (P = 0.012). As proline betaine is commonly interpreted as a biomarker of fruit intake, particularly citrus consumption, these findings may capture broader dietary influences on child behavioural development rather than a metabolite-specific effect. Additional negative associations were observed for leucine and N-acetylneuraminic acid with externalising symptoms, glycine with ADHD symptoms, and 5-oxoproline with internalising and total problems. Conversely, tyrosine, lysine and N^1^-methylnicotinamide were associated with higher behavioural problem scores across one or more domains.

**Figure 2.**
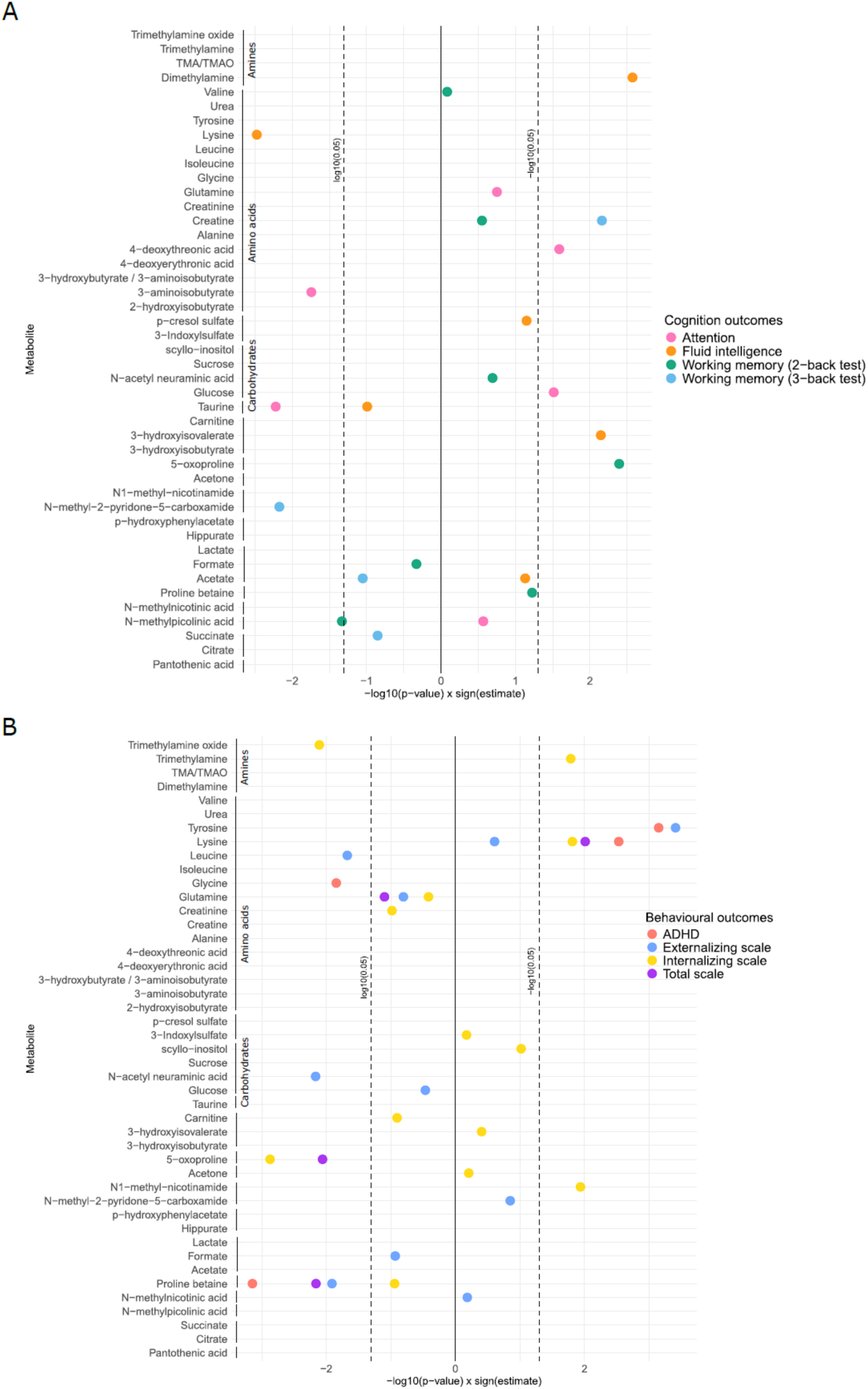
Urinary metabolite associations with neurodevelopmental outcomes in children of the HELIX subcohort (N=720-1203). ElasticNet models with SHARP (Stability-enHanced Approaches using Resampling Procedures) adjusting for maternal education, child’s age at assessment and cohort were fitted on 45 urine metabolites as measured by ^1^H-NMR spectroscopy to predict A) behavioural outcomes (N=1201-1203) and B) cognitive outcomes (N=720-989). Only the –log_10_ p-values for the metabolites that were selected by the models for each outcome are displayed. (A, B) Miami plots, the x-axes show the –log_10_ p-values multiplied by the direction of the association (sign of the regression coefficient) and the y-axes group metabolites; colours in panel legends indicate the outcome associated. For full details, see Table S1.

For cognitive outcomes, ElasticNet regression selected 19 of the 45 semi-quantified metabolites as predictive of at least one cognitive domain (Figure 2B). Outcome-specific multi-metabolite models including these selected metabolites identified several nominal associations with cognitive performance (unadjusted P < 0.05). Among methylamine-related metabolites, dimethylamine (DMA) was associated with higher fluid intelligence (P = 0.003), suggesting that methylamine metabolism may also be related to cognitive outcomes, although in a domain-specific manner. Several other metabolites showed positive associations with cognitive performance, including creatinine (working memory, P = 0.007), 4-deoxythreonic acid (attention, P = 0.026), 3-hydroxyisovalerate (fluid intelligence, P = 0.007), 5-oxoproline (working memory, P = 0.004) and glucose (attention, P = 0.031), whereas associations with lower cognitive performance were seen for lysine (fluid intelligence, P = 0.003), N-methyl-2-pyridone-5-carboxamide (working memory, P = 0.007), N-methylpicolinic acid (working memory, P = 0.047), 3-aminoisobutyrate (attention, P = 0.018) and taurine (taurine, P = 0.006).

Among the metabolites identified in the behavioural and cognitive screening analyses, the methylamines TMAO, TMA and DMA were of particular interest because they were selected across neurodevelopmental domains and showed divergent directions of association. TMAO was associated with lower internalising problems, TMA with higher internalising problems, and DMA with higher fluid intelligence. Because TMAO can reflect seafood intake^22^, we further assessed whether the association between TMAO and internalising problems was explained by fish intake. Adjustment for the serum sum of polyunsaturated fatty acids (PUFAs), a recognised biomarker of fish consumption^19^, did not alter the association. The TMAO estimate remained unchanged in the basic and PUFA-adjusted models (β = –4.10^-05^, 95% CI: [-7.10^-05^,-1.10^-05^] in both models), with similar nominal P values before and after PUFA adjustment (P = 0.0078 and P = 0.0075, respectively). These findings supported the case for an effect of TMAO independent of fish intake and prioritisation of methylamine metabolism for longitudinal and experimental follow-up.

### Prenatal exposure to TMAO, TMA and related amines associates with later neurodevelopmental outcomes

To test whether prenatal exposure to TMAO, TMA, and related metabolites (such as choline and DMA) were linked with neurodevelopmental outcomes, we assessed longitudinal associations between metabolites measured during pregnancy and neurodevelopment in children, aiming to disentangle potential reverse causation effects of diet or other unmeasured confounders of the association between maternal urinary TMAO and neurodevelopment. The INMA cohorts followed 815 mother–child pairs, assessing neurodevelopmental outcomes at ages 1, 4, 8, 9, 11, and 14 years. Repeated outcome measurements yielded more than 10,000 assessments over the course of the study, allowing us to examine longitudinal changes in cognitive and behavioural outcomes. TMAO, TMA, and related metabolites were semi-quantified through urinary ^1^H-NMR metabolomics in pregnant women in the first and third trimesters, with the same platform as used with the HELIX cohort data above. Linear mixed effects models were used adjusted by maternal education, season of birth, child’s age at assessment (different for each outcome and across timepoints), maternal age, parity, and cohort, with a random intercept for child.

Maternal urinary levels of TMA in the third trimester of pregnancy were significantly associated with an increased risk of externalising behaviour in children, after FDR correction (p_adj_ = 0.024) (Figure 3A). TMA measured in the first trimester was also nominally associated (p-value < 0.05, uncorrected) with lower non-verbal intelligence (p-value = 0.038) (Figure 3B). Overall, these results suggest that TMA exerted a detrimental influence on child neurodevelopment at all ages, supporting the previous cross-sectional results in childhood. However, prenatal TMAO exposure showed no association with beneficial long-term neurodevelopment. These results remained consistent when models were further adjusted for obstetric complications or maternal fish intake during pregnancy (Table S2). Finally, TMA associations in the first and third trimesters were observed predominantly in boys, when we modelled separately by sex, suggesting a greater detrimental effect of TMA in boys (Figure 4A-D).

**Figure 3.**
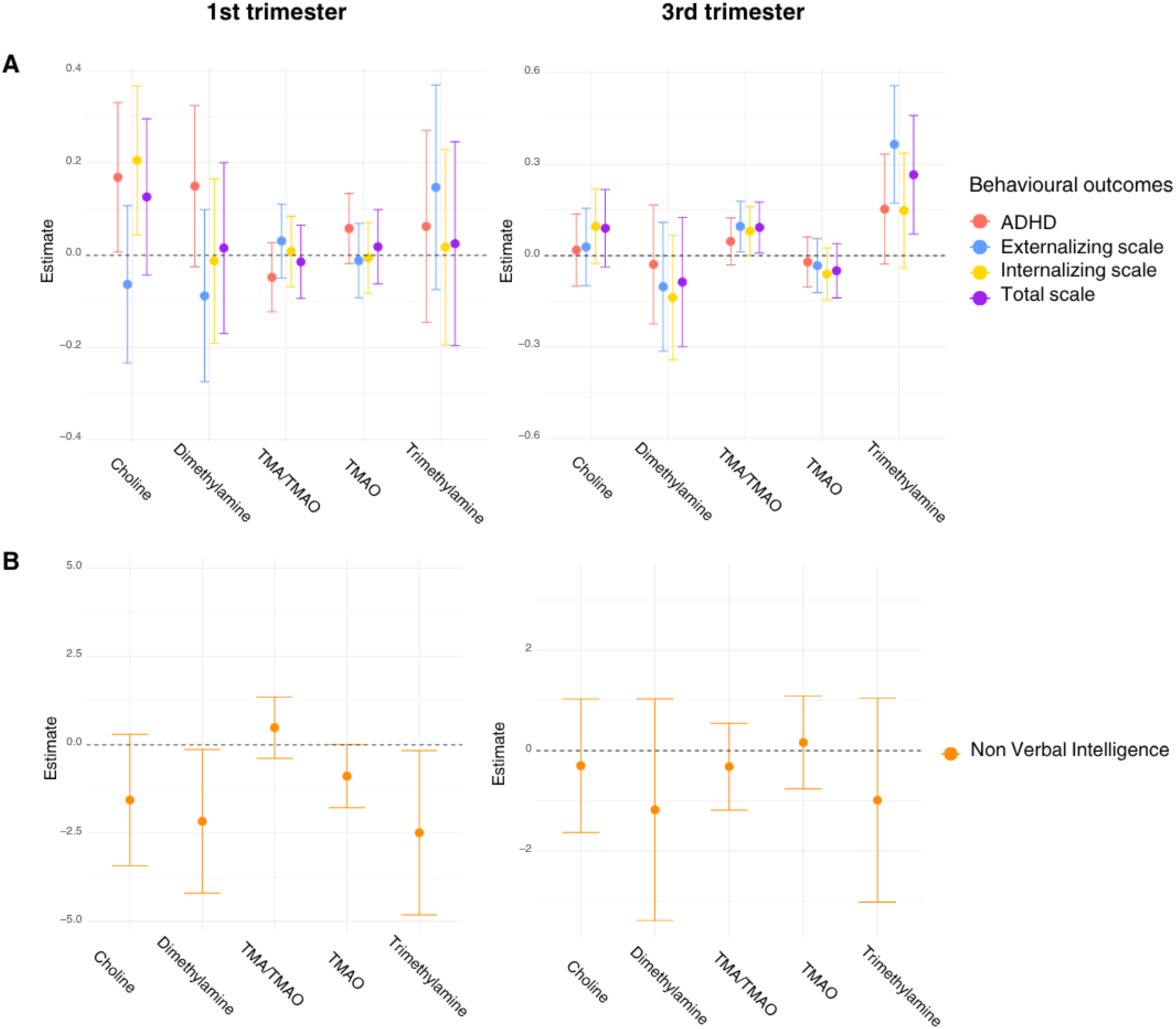
Prenatal maternal urinary TMAO and related metabolite associations with child and adolescent cognition and behaviour problems in the INMA cohorts. Linear mixed-effect regression models were computed between each first and third trimester metabolite and outcomes, adjusting for maternal education, season of birth, child’s age at assessment (different for each outcome and across timepoints), maternal age, parity and cohort, with a random intercept for child. The estimate on the y-axis is the estimated effect size (fixed-effect coefficient) from the linear mixed-effects model. A) Behavioural problems: ADHD symptoms, N = 668 (assessed at 7, 9, 11, and 14 years old, T = 1751 data points) and CBCL scales, N=630 (assessed at 8 and 15 years old, T = 1057 data points), B) Non-verbal intelligence, N= 820 children (assessed at 15 months, 4, 9, and 14 years old, T=2267 data points).

**Figure 4.**
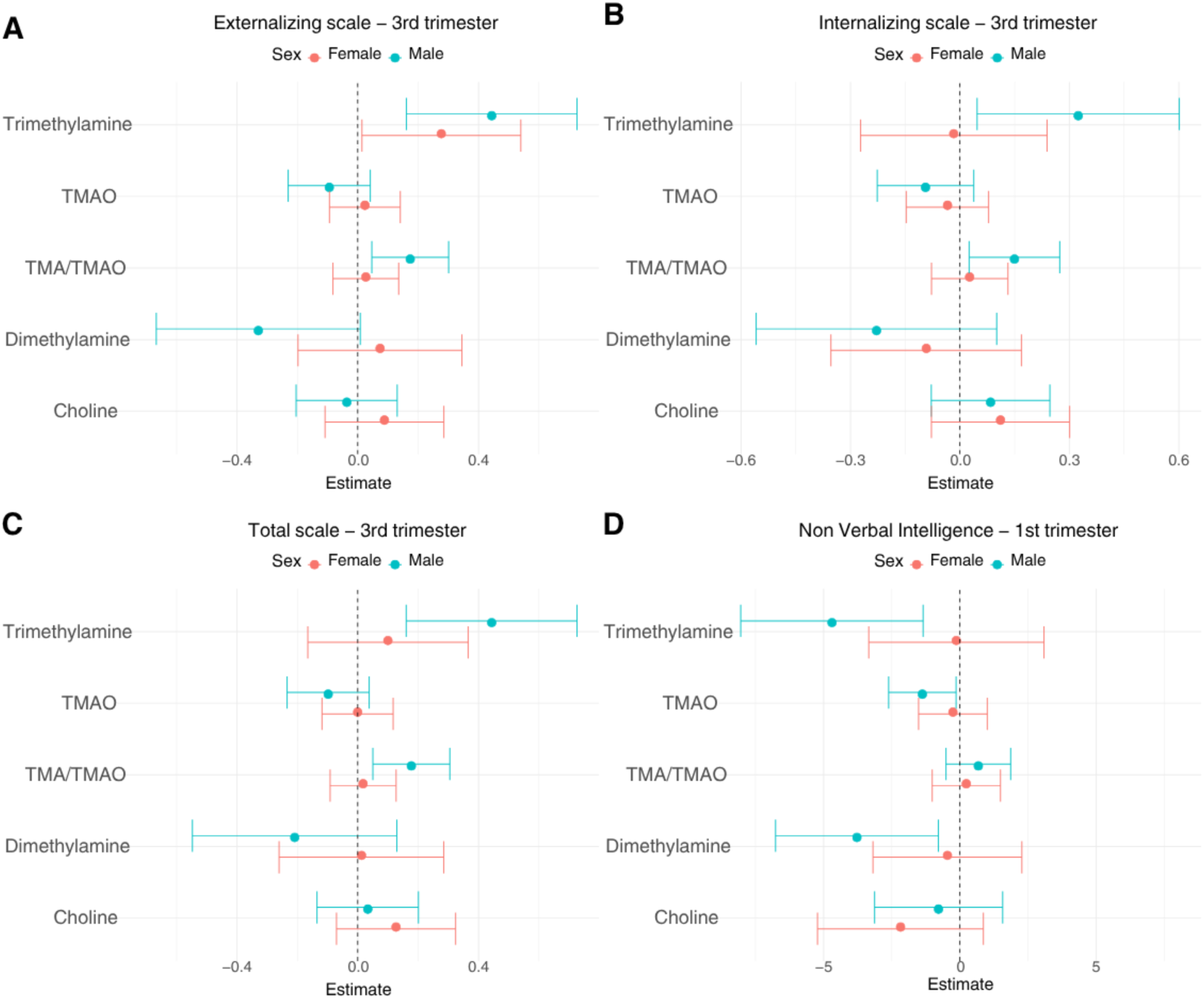
Sex-stratified associations between prenatal TMAO and related metabolites and childhood neurodevelopmental outcomes. Sex differences in associations between prenatal maternal urinary TMAO and related metabolite levels with A-C) behavioural problems and D) non-verbal intelligence in childhood and adolescence in the INMA cohort. Behavioural outcomes analyses included: females, N = 320 (T = 545 datapoints); males, N = 310 (T = 512 datapoints). Non-verbal intelligence analyses included: females, N = 410 (T = 1145 datapoints); males, N = 409 (T = 1121 datapoints).

TMAO and TMA regulate neuronal differentiation-related genes in differentiating stem cells Building on these associations from the HELIX and INMA cohorts, we wanted to investigate the biological plausibility of the potential adverse effects of TMA and beneficial effects of TMAO using mechanistic information. Previous research has shown exposure to TMAO during development to have a beneficial effect on axonogenesis and cognitive function in mice^20^ and to enhance barrier integrity in a human-cell-based model of the blood–brain barrier (BBB)^10^. To determine whether TMAO similarly affected human neurons, and how these cells might be affected by TMA, we employed an established human stem cell-derived model, the human neural progenitor test (hNPT)^21^. Neural progenitor cells (NPCs) were differentiated to neurons and astrocytes while being exposed to physiologically relevant, non-cytotoxic concentrations of TMAO or TMA (40 µM or 0.4 μM, respectively^22^) for 10 days (Figure 5A).

**Figure 5.**
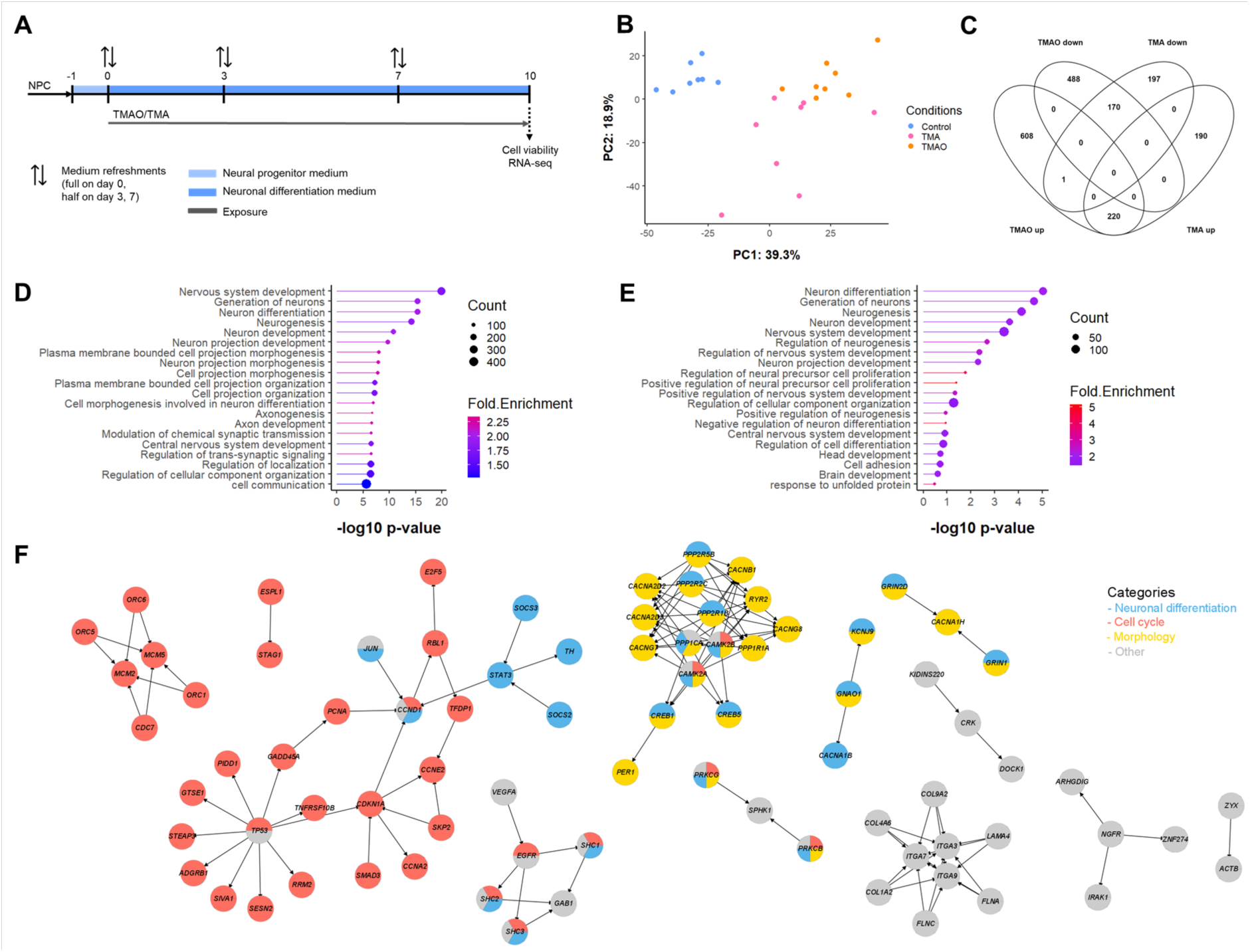
Effects of TMAO and TMA on neuronal–glial differentiation in the hNPT. A) Schematic representation of the hNPT. B) PCA scores plot of TMAO and TMA based on genes regulated by either of the compounds (p_FDR_ <0.05). C) Venn diagram of significantly up– and down-regulated genes by TMAO or TMA (p_FDR_ <0.05). D-E) Top enriched GO-terms by TMAO (D) and TMA (E). F) Network of regulated genes from the overrepresented pathways, colour-coded according to the group the pathway was assigned to. A list of all enriched GO-terms and enriched pathways can be found in Table S3 and Table S4, respectively. NPC, neural progenitor cell; TMAO, trimethylamine N-oxide; TMA, trimethylamine; PC, principal component. Data represents one independent experiment (N=1) with eight biological replicates (n=8).

Upon bulk RNAseq analysis, TMAO was found to have regulated expression of 1487 genes (829 up, 658 down; p_adj_<0.05) while TMA regulated 778 genes (410 up, 368 down; p_adj_<0.05). Principal component analysis (PCA) of these genes revealed distinct clustering of TMAO and TMA-exposed hNPT (Figure 5B). Interestingly, many genes that were regulated by TMAO or TMA followed the same direction seen with the other compound (Figure S1A) and all genes that were regulated by both methylamines were regulated in the same direction (Figure 5C), consistent with earlier findings^10^. Gene Ontology (GO) analysis of the regulated genes showed 86 GO-terms significantly enriched by TMAO and 11 by TMA (Table S3). Top enriched GO-terms of both compounds overlapped considerably, notably including GO-terms related to neuronal differentiation and cell morphology (Figure 5D, E). TMAO exposure additionally enriched GO-terms related to synaptic signalling and cell cycle that did not reach the top enriched GO-terms, but were clearly and statistically enriched compared to TMA (Table S3, Figure S1B, C).When genes uniquely regulated by TMAO were analysed, those GO terms related to synaptic signalling were significantly enriched.

Pathway analysis was undertaken for the TMAO or TMA-regulated genes, mapping expression of differentially expressed genes onto all human pathways in the KEGG database. Using Signalling Pathway Impact Analysis (SPIA), genes regulated by TMAO exposure were overrepresented in 11 pathways (1430 input genes, SPIA p_GFDR_<0.05), while TMA-regulated genes were not overrepresented on any pathway (746 input genes, SPIA p_GFDR_<0.3) (Figure S1D, E; Table S4). All overrepresented pathways were activated by TMAO, with the exception of the p53 signalling pathway, which was inhibited. The network of the 105 genes underlying these pathways revealed hub genes such as *TP53* and protein phosphatase 2 regulatory subunits (*PPP2R1B, PPP2R2C, PPP2R5B*) important for cell growth and division, and calcium/calmodulin-dependent protein kinase II A and B (*CAMK2A, CAMK2B*), which are important in calcium signalling and synaptic plasticity (Figure 5F, Figure S1F, Table S4). Activated pathways could be divided in three groups: cell cycle, neuronal differentiation and neuron/neurite morphology, in line with the GO-term analysis results (Figure 5F). These results together show that TMAO gene regulation specifically converged in pathways that point toward activation of these three groups, that would be associated with increased/more rapid maturation of neurons and increased neural network activity, while TMA did not.

We also compared the differentially expressed genes for TMAO in the hNPT to those seen upon similar treatment of the human cerebrovascular endothelial hCMEC/D3 cell line, a BBB cell model^10^. TMAO exposure in hNPT and hCMEC/D3 cells had 31 overlapping genes. These genes were mostly involved in cell cycle and cell morphology, suggesting a common effect of TMAO on two different types of cells in the brain (Figure S1G, Table S4).

Interaction between TMAO and environmental pollutants *in vitro* and in human cohorts reveals potential protective effect of TMAO

As described above, TMAO exposure co-occurs with environmental contaminants, most importantly Hg and As, due to the association of all three with fish consumption^16^. We therefore investigated the interaction of TMAO with these contaminants *in vitro* and in the HELIX and INMA cohorts.

We first examined gene expression in hNPT cultures following 10 days of exposure to either MeHg or NaAs at concentrations just below those causing a decrease in cell viability (IC_5_ – MeHg, 0.1 μM; NaAs, 1.2 μM) or one well below cytotoxic levels (IC_5_/100 – MeHg, 1 nM; NaAs, 12 nM) and within the range of physiological blood concentrations detected in European children (MeHg, 1.2–2.5 nM; As, 3.3–18.6 nM^23^) (Figure 6A, Figure S2A). Exposure to IC_5_ concentrations of MeHg or NaAs affected neuronal differentiation, synaptic signalling, cell cycle and morphology-related GO-terms (Table S3). An IC_5_/100 concentration of MeHg induced apoptosis-related GO-terms only, while NaAs at this concentration did not enrich any GO-terms. SPIA revealed that, at the pathway level, the observed effects were mostly in the opposite direction to those induced by TMAO for an IC_5_ concentration of MeHg and NaAs, and an IC_5_/100 concentration of MeHg (Figure S2C-H, Table S4).

**Figure 6.**
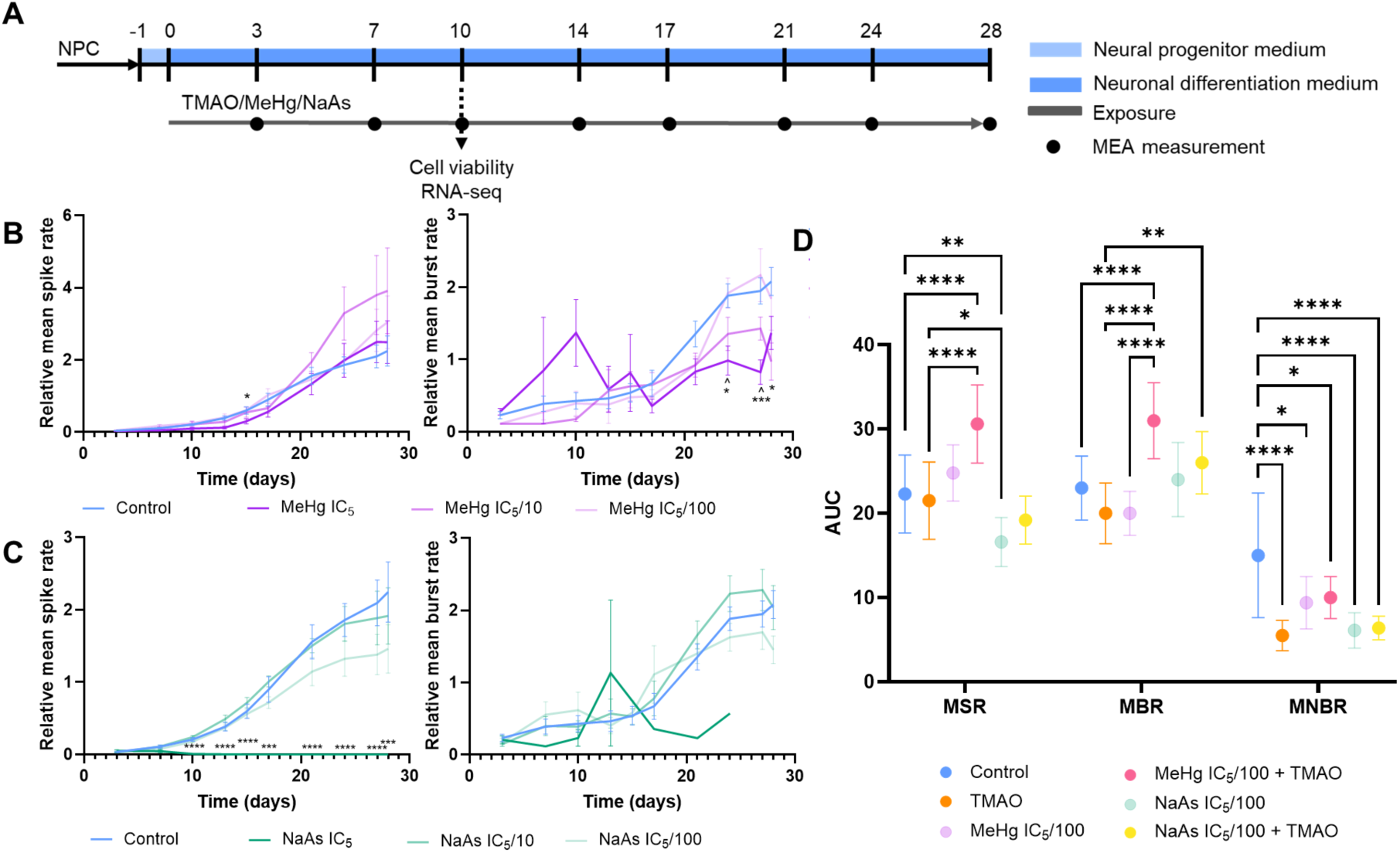
Neuronal network formation is modulated by TMAO, MeHg or NaAs. A) Culture and exposure protocol of the hNPT. B,C) Neuronal network formation was measured as spontaneous electrical activity in the hNPT. Relative mean spike rate (MSR) and mean burst rate (MBR) over time in the hNPT when exposed to either MeHg or NaAs at different concentrations For NaAs MSR, only significance relative to the control was plotted. *, significantly different to control; ^, significantly different to IC_5_/100. D) Area under the curve of developmental trajectories of different exposure conditions. The number of * in B-D represent * p<0.05, ** p<0.01, *** p<0.001, *** p<0.0001, N=2, n control = 20, n TMAO = 17, n NaAs IC_5_, IC_5_/10, IC_5_/100 = 11, n NaAs IC_5_/100 + TMAO = 10, n MeHg IC_5_ = 9, n MeHg IC_5_/10, MeHg IC_5_/100 + TMAO = 8, n MeHg IC_5_/100 = 7.

We next employed an extended treatment protocol to study functional effects of MeHg or NaAs, in which hNPT cultures were exposed for 4 weeks to various concentrations of MeHg or NaAs (Figure 6A). Exposure to either MeHg or NaAs dose-dependently impaired neuronal network formation, with NaAs being the more potently detrimental. While the highest concentration of MeHg significantly affected the relative mean burst rate (MBR) in the cultures, it had no effect on the relative mean spike rate (MSR), suggesting a disruption in network formation (Figure 6B)^24^. In contrast, NaAs effects were most apparent in the relative MSR and only at the highest concentration, i.e. when cell viability was decreased by 5 %. At this point the spike rate dropped to almost zero, clearly affecting the individual neurons rather than network formation alone (Figure 6C). Co-treatment with 40 μM TMAO and a physiologically relevant concentration of MeHg (IC_5_/100, 1 nM) caused a significant increase of the relative MBR (p < 0.0001) and non-significantly increased the relative MSR (p = 0.07). Interestingly, TMAO did not have this effect on its own, nor did it mitigate the effects of a physiological concentration of NaAs (IC_5_/100, 12 nM) (Figure 6D). The relative mean network burst rate (MNBR) was not changed with either compound. These data indicate that it is biologically plausible that TMAO can mitigate against the negative effects of some neurodevelopmental toxicants such as Hg, *via* the promotion of neuronal network formation.

In the HELIX and INMA cohorts, we also had information on total Hg and As concentrations in whole blood of children and in urine of pregnant mothers. Associations between As and fluid intelligence depended on TMAO levels in childhood, but not in pregnancy. Specifically, the association between As exposure in childhood and fluid intelligence differed across TMAO tertiles. Among children in the lowest TMAO tertile, As was negatively associated with fluid intelligence (estimate: –0.35; 95% confidence interval (CI): [-0.82, 0.13]), whereas positive associations were observed in the medium (estimate: 0.36; 95% CI: [-0.06, 0.79]) and high tertiles (estimate: 0.46; 95% CI: [0.08, 0.84]) (Figure 7A). Formal interaction testing confirmed significant effect modification by TMAO levels (Likelihood Ratio Test p-value < 0.05). A similar trend was observed for Hg exposure, with negative effects on fluid intelligence only observed in the children within the lowest TMAO tertile (estimate: –0.69; 95% CI: [-1.39, 0]), while no associations were observed within the higher TMAO tertiles. This result suggests that TMAO modifies the association between neurotoxic metals and cognitive function. TMAO in pregnancy did not have the same effect in the stratified analysis in the longitudinal INMA cohorts, when the exposure and TMAO were measured during pregnancy (Figure 7B). Interactions are shown for all outcomes in Table S1-8,S1-9 and S2-8.

**Figure 7.**
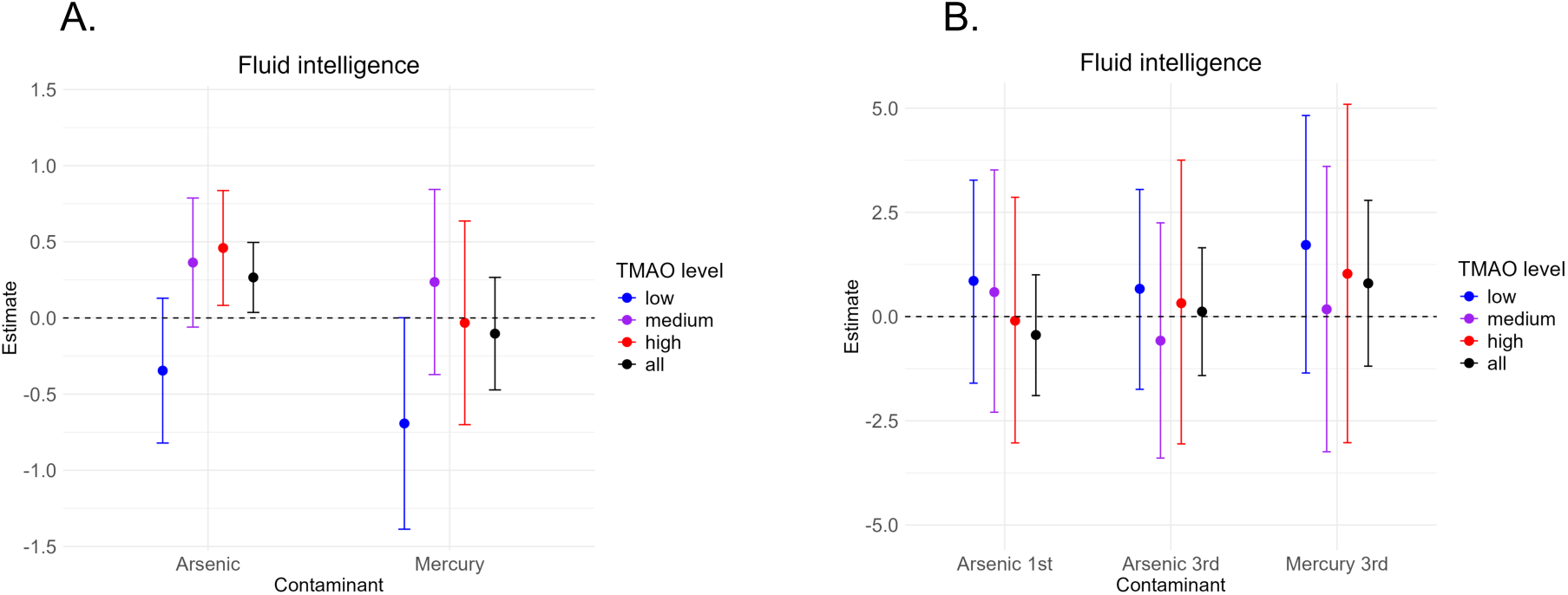
Associations between As and Hg exposures with cognitive function in children. Exposures were determined in A) childhood cross-sectionally or B) during pregnancy. To determine potential interaction by levels of TMAO, analyses were stratified for children with different levels of TMAO (tertiles) in their urine at childhood (A) or the urine of their mothers in the first and third trimester (B). N = 989 (cross-sectional) and N = 385-675 (during pregnancy, As first trimester: 385, As third trimester: 441, Hg third trimester: 675).

## Discussion

Here, we report that the MHCM TMAO is associated with more favourable neurodevelopmental outcomes in children, with *in vitro* analyses suggesting that this may involve enhancement of neuronal differentiation pathways. In contrast, TMAO’s microbially-produced precursor TMA is consistently associated with behavioural problems in childhood, and prenatal TMA exposure with worse childhood neurodevelopmental outcomes. TMAO also appeared to attenuate adverse associations between environmental metal contaminants and childhood neurodevelopment and protected against MeHg-induced changes in neuronal function *in vitro*. These results reinforce the concept that MHCMs can modulate human neurodevelopment, confirming the utility and power of combining population-scale exposome studies with cellular toxicology to unravel human biology based on mechanistic insights with *in vitro modelling*.

Very few studies have looked at TMAO and child neurodevelopment. Two studies in children found higher TMAO to be associated with higher visual memory scores^25^, and improved developmental progress^26^. However, the latter study did not correct for fish intake, making it hard to interpret as circulating TMAO can arise from both direct dietary intake and from host–microbial co-metabolism; DMA and TMA can be generated through microbial catabolism of dietary choline, TMAO, L-carnitine, phosphatidylcholine and betaine in the gastrointestinal tract. After absorption, TMA is transported to the liver, where it is oxidised by hepatic flavin-containing monooxygenases to TMAO^20,21^. TMAO is also abundant in seafood^22^, and a proportion of ingested TMAO can undergo microbial reduction to TMA^20,23^ before retroconversion to TMAO by host enzymes^20,23^. Thus, circulating TMAO may reflect both seafood intake and microbial methylamine metabolism. In our study, TMAO was associated with beneficial effects on behaviour in the cross-sectional cohort study, and this association was not altered after adjustment for serum PUFAs, a biomarker of fish intake. At the cellular level, beyond its previously reported tightening effect on the BBB^19^, TMAO promoted gene-expression changes consistent with neuronal and astrocytic development in differentiating human neural progenitor cells. Although TMAO did not cause major changes in neuronal network activity, transcriptomic analyses showed activation of neurodevelopmentally relevant pathways in the hNPT model, extending murine data showing that TMAO improves thalamic explant axonogenesis^24^ to human systems. These results add to growing evidence that TMAO may not be uniformly harmful for neurodevelopment and may even be beneficial at physiologically relevant concentrations.

To our knowledge, this is the first report of a role for TMA in neurodevelopment, identifying a consistent association between TMA and behavioural problems in humans. Gene expression analysis in the hNPT did not offer mechanistic explanations for these strong epidemiological findings, however. If anything, there seemed to be a lack of effect of TMA, perhaps indicating a mechanism outside of direct interactions with neuronal or astroglial cell types found in the hNPT. We have previously shown TMA to negatively affect the BBB in a human cell line model^10^, suggesting that it may have rendered the developing brain more accessible to exogenous compounds^10^. This is not to say that TMA is necessarily detrimental in all circumstances, as we have also recently shown beneficial effects of the molecule on metabolic inflammation^27^, again emphasising the importance of biological context in interpreting the effects of MHCMs and their contributions to the endogenous exposome. Therefore, a single *in vitro* model is not sufficient to study the effect(s) of TMA on brain development. It can only be concluded that TMA does not influence the mode of action related to neuron–astrocyte differentiation.

A notable feature of the data is the sex-specific nature of many of the TMA associations, with effects predominantly seen in males. While the majority of children studied will have been pre-pubertal given their age, it is important to note that this does not necessarily preclude an interaction between TMA and sex hormones. Sexual differentiation of the brain is driven in large part by the organisational effects of the male-specific testosterone surge that occurs during the second trimester^28,29^. While our measurements were taken in the first and third trimesters, it seems plausible that in many cases TMA levels may be comparable during the second trimester. Assuming this, these results suggest a possible interaction between TMA and the sexually differentiating brain environment. Additionally, adult males have been reported as expressing lower levels of hepatic FMO3 compared with females, resulting in higher circulating levels of TMA^30^. It is unknown whether similar sex differences exist prenatally, as hepatic FMO3 expression is very low during prenatal development^31^, but this may be an additional source of sex differences. Furthermore, sex differentiation of the brain is not reflected in our hNPT, reflecting that this model does not represent TMA’s mode of action with respect to brain development.

Extending the evidence for the potentially beneficial role of TMAO, stratification of the cross-sectional data showed that the effect of As exposure in childhood and fluid intelligence depended on TMAO levels, i.e. low TMAO levels resulted in lower fluid intelligence compared to high, protective TMAO levels. *In vitro*, TMAO could diminish negative effects of an environmentally relevant concentration of MeHg on neural network activity, although notably it was ineffective against NaAs toxicity. This may reflect the varying mechanisms through which MeHg and NaAs are thought to exert their toxic effects, inhibition of neuronal migration and disruption of cell energy balance for MeHg^32^ and induction of ROS formation and apoptosis for NaAs^33^. Notably, TMAO has been shown to interact with mitochondrial function^34,35^ and hence energy balance, but building a full understanding of the interactions of this MHCM and these environmental toxicants remains a subject for future study. It is plausible that there are other neuroprotective compounds in fish that co-occur with TMAO and are protective against Hg or As exposure, most obviously including polyunsaturated fatty acids. Indeed, our analysis of the HELIX cohort data identifies beneficial associations with cognition for several metabolites known to be obtainable from fish, e.g. 5-oxoproline and 4-deoxythreonic acid. Detailed exploration of all possible interactions lies outside the scope of a single study however. Future analysis of more complex *in vitro* models, e.g. inclusion of additional CNS cell types in the hNPT, or use of multi-system organ-on-a-chip platforms may also provide insights in the complex interface between MHCMs, toxins and neurodevelopment.

It should also be noted that for the longitudinal epidemiological analyses, while we adjusted for several relevant confounders such as fish intake, residual confounding remains possible. This is especially the case given the extended time (up to 15 years) between prenatal metabolite exposure and assessment of neurodevelopmental outcomes in childhood, during which additional relevant factors may not have been fully captured. Nonetheless, the fact that signatures of prenatal MHCM exposure could be detected in childhood neurodevelopmental outcomes even at this distance attests to the significance of these associations and corroborate studies in mice^20^.

Taken together, our data emphasise the context-dependent nature of MHCMs and their effects on the body, i.e. the relative beneficial/detrimental actions of given MHCMs are intricately bound up with the physiological status of the host. As an example, TMAO was initially linked to an increased risk of preeclampsia^36–38^, which was later contradicted once dietary fish intake was taken into account^39^. Given that the primary variable predicting circulating TMAO levels is kidney function^40^, more complex, longitudinal analyses incorporating a much broader range of physiological, dietary and microbial parameters are needed to truly understand the role of TMAO (and other MHCMs) in mammalian biological processes. Combining this with human-based experimental models is also crucial to increase translational relevance; this is, for example, particularly relevant in the case of TMA, which is present at more than ten times higher levels in the urine of mice than humans^41^.

## Conclusions

Links between the gut microbiota and cognition are increasingly being identified, but in general the underlying mechanistic pathways are less well understood. Here, we have focused on an exemplar family of MHCMs, the methylamines, employing a combination of population studies and *in vitro* models to examine their impact upon neurodevelopment. We have found significant associations with neurodevelopmental outcomes in children, with a notable distinction between the detrimental effects of TMA and the more positive actions of its metabolite TMAO. Moreover, TMAO levels appear to modify the association between cognitive function and exposure to the environmental neurotoxicants Hg and As, with negative effects of these pollutants only observed when TMAO levels are low. Notably, in the case of Hg, these effects could be replicated *in vitro* by TMAO-induced upregulation of neuronal differentiation pathways and improved neuronal network function. Taken together, these studies demonstrate how advances in epidemiology, specifically exposome studies of a healthy, population-based birth cohort, can be combined with state-of-the-art human-based model systems to gain mechanistic insights into the effect of environmental exposures on humans and how the gut microbiota, *via* metabolic output, might influence the developing brain.

## Data Availability

All data produced in the present study are available upon reasonable request to the authors

https://doi.org/10.6084/m9.figshare.31076257

https://github.com/HERMES-Exposome/TMAO_exposure_neurodevelopment.git

## Materials & Methods

### Studied population

The study population is based on the HELIX (Human Early-Life Exposome) project, which pooled data on existing six population-based birth cohorts (BiB in United Kingdom, EDEN in France, KANC in Lithuania, INMA in Spain, MoBa in Norway, and RHEA in Greece) where large sets of previously collected longitudinal data from early pregnancy to childhood were available. Background information on the HELIX cohorts and the full HELIX protocols are described elsewhere^42–44^. The HELIX subcohort consists of 1,301 mother child–pairs that were followed-up at 6–11 years of age using common protocols for all the cohorts (between 12/2013 and 02/2016) including biological samples collection, face-to-face questionnaire, health examination and additional characterisation for a wide range of exposures^45^. The final sample size for the cross-sectional analyses in children, included 1,203 children with at least one outcome data available and urine metabolites.

INMA (INfancia y Medio Ambiente) is a birth cohort study in seven regions of Spain that aims to examine the role of environmental pollutants in relation to child growth and development. All participants were singleton live-born infants from two INMA subcohorts located in Gipuzkoa (Basque Country) and Sabadell (Catalonia) [12]. The women were interviewed twice during pregnancy (in the first and third trimesters of gestation) to obtain information about their sociodemographic characteristics and lifestyle variables. Urine samples were collected in the same interview in the morning (spot samples). Urine was collected in 100 mL polyethylene containers and stored at −20 °C. One aliquot of the sample from each of the participants was sent to the laboratories of the Department of Surgery and Cancer, Imperial College London, UK, to be analysed. NMR spectra of urine were generated from 412 and 417 subjects for the first and third trimesters, respectively (12.4 ± 1.2 and 33.9 ± 1.3 weeks, respectively) in Gipuzkoa and from 394 and 469 subjects for Sabadell.

The study was approved by the individual national ethics committees for cohort recruitment, follow-up visits and secondary use of pre-existing data for HELIX. Written informed consent was obtained from all HELIX subcohort participants.The INMA project was approved by the Ethical Committees of the participating centres, and all subjects gave written consent at enrolment and delivery.

### Cognition, emotional and behavioural problem assessments

Cognitive, emotional and behavioural outcomes were assessed in children from the HELIX cohort and in the INMA Sabadell and Gipuzkoa cohorts using standardised neuropsychological tests and parent-reported questionnaires. In HELIX, parents completed questionnaires related to the child’s behavior, including the Conner rating scale’s (N = 1287) and child behavior checklist (CBCL, N = 1298), within a week before the follow-up visit at 6–11 years of age. The 99-item CBCL/6–18 version for school children was used to obtain standardized parent reports of children’s problem behaviours, translated and validated in each native language of the participating six cohort populations^46^. The parents responded along a 3-point scale with the code of 0 if the item is not true of the child, 1 for sometimes true, and 2 for often true. The internalising score includes the subscales of emotionally reactive and anxious/depressed symptoms, as well as somatic complaints and symptoms of being withdrawn. The externalising score includes attention problems and aggressive behaviors. In addition, an ADHD index based on the short form of the Conners’ rating scales of 27 items provided information on inattention and hyperactivity symptoms^47^. The internal consistency (Cronbach’s alpha) of each of the study scales was >0.80. All outcomes were analysed as raw count scores, which were square root-transformed and subsequently z-standardised.

Trained fieldwork technicians measured three cognitive domains in children using a battery of computer-based tests: fluid intelligence (Raven Coloured Progressive Matrices Test [CPM]), attention function (Attention Network Test [ANT]) and working memory (N-Back task). The CPM comprised a total of 36 items and we used the total number of correct responses as the outcome. A higher CPM scoring indicates better fluid intelligence. Fluid intelligence is the ability to solve novel reasoning problems and depends only minimally on prior learning^48^. For ANT, we used the outcome of hit reaction time standard error (HRT-SE), a measure of response speed consistency throughout the test. A high HRT-SE indicates highly variable reaction time during the attention task and is considered a measure of inattentiveness^49^. As the main parameter of N-Back, we used d prime (d′), a measure derived from signal detection theory calculated by subtracting the z-score of the false alarm rate from the z-score of the hit rate. A higher d′ indicates more accurate test performance, i.e. better working memory^49^. The N-Back task was also completed by the mothers. All examiners were previously trained following a standardised assessment protocol by the study expert psychologist. Furthermore, during the pilot phase, a coordinator visited each cohort site and checked for any potential error committed by the previously trained examiners. We used all the outcomes as cognitive gross scores and we adjusted them by child age in the regression models. Complete outcome descriptions are provided in previous publications^43,50^.

In INMA Sabadell and Gipuzkoa, neurodevelopmental outcomes were assessed longitudinally from infancy to adolescence, with repeated measures available at approximately 1, 4, 8, 9, 11, and 14 years of age depending on the cohort and follow-up wave as described previously. Early cognitive development was assessed at a mean age of 1.2 years using the Bayley test and at a mean age of 4.5 years using the McCarthy test. For both tests, early mental and motor scales were used, with higher scores indicating better performance. At later ages, INMA assessed cognitive domains comparable to HELIX using the same or closely related tests. Attention function was assessed at a mean age of 4.5 years using the Conners’ Kiddie Continuous Performance Test, a shorter test adapted for younger children, and at mean ages of 7.3, 9.1, 11 and 15 years using the same ANT as in HELIX. The selected outcome was hit reaction time standard error, with lower scores indicating better attention performance. Working memory was assessed using the same N-back task as in HELIX at mean ages of 7.3, 9.1, 11 and 15 years. The selected outcomes were d prime scores for the 2-back and 3-back conditions, with higher scores indicating better performance. Fluid intelligence was assessed using Raven’s Progressive Matrices, as in HELIX, at mean ages of 8.6 and 16 years. The total number of correct responses was used as the outcome, with higher scores indicating better performance.

Behavioural outcomes in INMA were assessed using parent-reported questionnaires comparable to those used in HELIX. ADHD symptoms were assessed at mean ages of 7.2, 9.1, 11 and 14 years, with higher scores indicating more symptoms. Emotional and behavioural problems were assessed using the CBCL at mean ages of 8.5 and 15 years. Externalising, internalising and total problem scores were used, with higher scores indicating more behavioural or emotional problems.

### Urinary metabolic profiles

Urinary metabolic profiles were analysed by ^1^H-NMR on a 14.1 Tesla (600 MHz 1H) NMR spectrometer at Imperial College London, following a non-targeted approach. A total of 44 metabolites belonging to 22 metabolic classes were annotated: quantification was achieved for 24 metabolites, and the remaining 20 metabolites were semi-quantified as previously described^51,52^. Urine metabolite levels were normalised with the median fold change method and one-half of the minimum value was used as an offset. Then data were log_2_ transformed. A summary of urinary metabolite concentrations across the six cohorts can be found in Supplementary Material, Table S2 in Calvo-Serra et al. (2021)^53^.

### Arsenic and mercury measurements

In the HELIX cohort concentrations of metals in whole blood were measured at ALS Scandinavia (Sweden) using quadrupole inductively coupled plasma mass spectrometry (Q-ICP-MS) according to Rodushkin et al. (2000), with a limit of detection (LOD) ranging from 0.003–3.03 µg/L. Both arsenic (As) and mercury (Hg) were measured in whole blood collected during the childhood follow-up visit (ages 6–11 years). In the INMA Sabadell and Gipuzkoa subcohorts, total urinary arsenic was quantified in spot urine samples collected at the first (12.4 ± 1.2 weeks) and third (33.9 ± 1.3 weeks) trimesters of pregnancy, alongside a panel of 22 other metals and metalloids. Urinary metal concentrations were normalised to creatinine concentration (measured by standard clinical chemistry assay) to correct for urine dilution. Mercury was measured in cord blood samples collected at birth. Full details of the exposure assessment methods for the INMA subcohorts are described in Maitre et al. (2018)^17^.

### Statistical analysis

#### Epidemiological associations between childhood urinary metabolites (cross-sectional) and child behavioural problems and cognition

Due to the skewness of the urine metabolites distribution, log_2_ transformations were applied. Additionally, the ratio TMA/TMAO was created. If zero values existed their values were replaced by the minimum value divided by 2. Then, the ADHD index and the CBCL behavioural problem scores outcomes were transformed, first with a square root and then converted into standardised z-scores. No transformation was needed for the cognitive outcomes.

We first systematically tested the epidemiological associations between each of the childhood urinary metabolites and each neurodevelopmental outcome, successively and independently, using single linear regressions adjusted considering the following potential main confounders: maternal education, child age, and the cohort. Further adjusted models were explored by additionally adjusting for PUFA sum serum. Neurodevelopmental outcomes included working memory, attention, fluid intelligence and child’s behaviour (externalising, internalising and total CBCL scales, and ADHD symptoms score).

For all urine metabolites, the effect size is reported as the log2 fold change (log2FC) in the outcome scores. Associations are interpreted such that, for a one-unit increase in urine metabolite levels, the outcome is expected to change by the estimated regression coefficient on average. Multiple testing p-value correction was applied for each metabolite and outcome using the Benjamimi–Hochberg method, controlling for False Discovery Rate (FDR).

Second, a multi-exposure linear regression model approach was employed by allowing multiple metabolites included in the model. For this purpose, a variable selection algorithm ElasticNet was used (R package *glmnet*), allowing forcing confounders in the model combined with a resampling procedure method implemented in the R package sharp (Stability-enHanced Approaches using Resampling Procedures) which provides an integrated framework for stability-enhanced variable selection, graphical modelling and clustering. In stability selection, a feature selection algorithm was combined with a resampling technique to estimate feature selection probabilities. Then, for each outcome, stratified analyses by sex were conducted.

Finally, to explore the potential mediating effect of TMAO on the association between contaminants and neurodevelopmental outcomes, we used linear regression models to study the association between As, Hg and each outcome, stratifying the analysis by tertiles of TMAO. Additionally, we conducted interaction analyses between TMAO levels and each contaminant. For each outcome, we used a first model including the contaminant, TMAO tertiles, all confounders, and an interaction term between TMAO tertiles and the contaminant; and a second model without the interaction term. Then the significance of the interaction was assessed using a Likelihood Ratio Test (LRT) between the two models, implemented with the *lrtest* function and a p-value threshold of 0.05.

#### Longitudinal associations between prenatal gut microbial (TMAO, TMA, ratio TMA/TMAO, choline and DMA) urine metabolites with cognitive and behavioural trajectories in early and late childhood

To assess the associations of urine metabolites with cognitive, motor and behavioural trajectories, log2 transformations were applied to the exposures and transformations were considered for the outcome variables. Again, the ADHD index and the CBCL behavioural problem scores outcomes were transformed first with a square root and then converted into standardised z-scores. Then, non-verbal intelligence and motor skills scores were centred to 100 with a standard deviation of 15. No transformation was applied to working memory and inattention measures.

Linear mixed effects models adjusted by maternal education, season of birth, child’s age at assessment (different for each outcome and across timepoints), maternal age, parity and cohort, with a random intercept for child, were employed for the main adjusted longitudinal analyses. For running the linear mixed effects models we used the R package *lmer4* and p-values were adjusted using the FDR method. Further adjusted models were explored considering adding one variable at a time to the main adjusted model: Obstetric complications (birth weight < 2500 g or > 4000 g or gestational age <37 weeks) and total fish intake (trimester specific metabolites). Stratified analyses by sex were conducted as well.

Finally, as previously described in the cross-sectional section, we conducted a similar analysis for the contaminants, this time employing linear mixed effect models. We first examined the associations between contaminants (arsenic measured in the first and third trimesters of pregnancy and mercury in the third trimester) and neurodevelopmental outcomes, stratifying by tertile of TMAO or TMA. Then we performed an interaction analysis, comparing models with or without an interaction term between TMAO (or TMA) and the contaminant. The significance of the interaction was assessed using a Likelihood Ratio Test (LRT) between the two models, implemented with the *lrtest* function and a p-value threshold of 0.05.

### Cell culture and exposures

The cell culture was generated from a two-step protocol, as described before^54^. Female H9 human embryonic stem cells (WA09, RRID:CVCL_9773, passage 56, WiCell, Madison, WI, USA) were differentiated into neural progenitor cells (NPCs) and stored in liquid nitrogen until use. Cells were screened for mycoplasma infections once per year. To start an experiment, NPCs (passage 1) were thawed and cultured as described in the neuronal induction protocol from Stemcell Technologies (#28782, Stemcell Technologies, Vancouver, Canada). Cells were kept in STEMdiff™ Neural Progenitor medium (Stemcell Technologies) on Poly-L-Ornithine (PLO, 15 µg/ml, Sigma-Aldrich, Saint Louis, MO, USA) – laminin (10 µg/ml, Sigma-Aldrich) coated 6-well plates (Corning, New York, NY, USA) in a humidified chamber (37 °C, 5% CO_2_, 3% O_2_). Each day, whole medium changes were performed till the NPCs reached 100% confluent, generally within seven days. Then differentiation to a neuron-astrocyte co-culture was initiated by dissociating and seeding the NPCs at 2.56 × 10^5^ cells/cm^2^ on 96-well plates for cell viability experiments (Greiner Bio-One, Kremsmünster, Austria), 24-well plates for RNA-seq (Corning), or PLO-laminin-coated 8-well micro-slides for immunostainings (Ibidi, Gräfelfing, Germany). Cells were kept in Neural Progenitor medium for one day, and for the rest of the experiment in differentiation medium based on a medium formulation from Gunhanlar et al.^55^: neurobasal medium, 10 µl/ml N2 supplement (100x), 20 µl/mL B-27 without retinoic acid supplement (50x), 10 µl/ml 5000 IU/ml Penicillin / 5000 µg/mL Streptomycin, 10 µl/ml nonessential amino acids (100x), 20 ng/mL recombinant glial cell line-derived neurotrophic factor (GDNF) and 20 ng/ml recombinant brain-derived neurotrophic factor (BDNF; all from Gibco, Waltham, MA, USA), 1 μM dibutyryl cyclic adenosine monophosphate (Sigma-Aldrich), 200 μM ascorbic acid (Sigma-Aldrich) and 2 μg/mL laminin (Sigma-Aldrich). For the MEA experiments, an adjusted differentiation medium was used to enhance neuronal activity, which consisted of BrainPhys neuronal culture medium (Stemcell Technologies),20 µl/ml B-27 Plus supplement (50x), 10 µl/mL 5000 IU/ml Penicillin / 5000 µg/ml Streptomycin, 1 μg/ml laminin, and for the first week supplemented with 20 ng/ml GDNF and 20 ng/ml BDNF. Half of the medium was refreshed every three to four days. On day 0 and 3, 10 ng/ml recombinant ciliary neurotrophic factor (CNTF; Gibco) was present in the medium to stimulate astrocyte differentiation. GDNF, BDNF, ascorbic acid, dibutyryl cyclic adenosine monophosphate and CNTF were all dissolved in sterile solutions.

For each toxicant exposure, solutions were freshly prepared. Trimethylamine *N*-oxide (TMAO, CAS# 1184-78-7, 95%), trimethylamine (TMA, CAS# 75-50-3, 25 wt % in H_2_O) and sodium (meta)arsenite (NaAs, CAS# 7784-46-5, ≥90% purity) were dissolved in medium, methyl mercury (II) chloride (MeHg, CAS# 115-09-3, Cl ∼13%) was dissolved in dimethyl sulfoxide (DMSO, CAS# 67-68-5; all from Sigma-Aldrich). Final concentration of DMSO in the medium of all experimental conditions was 0.25% (v/v), which did not affect cell viability or differentiation. All solutions were prepared in polypropylene vials.

Cells were exposed from day 0 of the neuronal-astroglial differentiation. Cells received the same concentration of a compound on day 0 (full medium refreshment) as on all the other refreshment days (half medium refreshment). For cell viability measurements and determining concentrations for follow-up experiments, three experiments with three technical replicates were performed for each experiment to construct concentration-response curves. On day 10, cell viability was measured by incubating the cells for 1 hour with Cell Proliferation Reagent WST-1 (Roche, Mannheim, Germany) and measured on a Spectramax® M2 spectrofluorometer (Molecular Devices, Berkshire, United Kingdom) at 544Ex/59ROAST software (version 70.3)^56^ in R (version 4.3.0)^57^ was used to produce concentration-response curves and deriving effect concentrations with a 95% confidence interval, using the exponential fitting model. Derived concentrations were used for follow-up RNAseq and MEA experiments.

### Gene expression analyses

#### RNA isolation

Samples were collected and processed as described before^21^. Briefly, for each experimental condition, eight replicates from a single experiment were fixed in QIAzol (Qiagen, Hilden, Germany) on day 10. Working with eight wells at a time, medium was replaced for QIAzol and cells were resuspended by pipetting forcefully, transferred to vials and put on ice. Vials were stored at –80 °C until further processing. Whole RNA extraction was performed according to Qiagen’s RNeasy® mini kit protocol, including the DNase digestion step. Concentration of extracted RNA was determined on the Qubit3 (Invitrogen, Carlsbad, CA, USA) and RNA quality was measured using the 2100 Bioanalyzer (Agilent Technologies, Amstelveen, the Netherlands).

#### RNAseq and downstream analyses

A summary of RNAseq methodological information can be found in Table S5. RNA samples were analysed using the TruSeq Stranded mRNA protocol (Illumina, San Diego, CA, USA). mRNA enrichment was performed by polyA-affinity purification, converting it into multiplex libraries of 16 samples for each flow cell. The NextSeq 500 sequencer (Illumina) using the NextSeq 500/550 High Output Kit version 2.5 (75 cycles) was used to sequence the samples. Raw bcl files were base called and demultiplexed into FASTQ files with Bcl2fastq (Illumina, version 2.20.0.422). Quality control reports (FastQC, version 0.11.5) were generated using MultiQC (version 1.0)^58^ to identify potential anomalies regarding library size, read length distribution, mean read quality distribution, mean quality for each position in the read, and base frequency for each position in the read. No aberrations were found in this process.

Next, FASTQ reads were mapped to the human reference genome (genome assembly: GRCh38, version 30, Ensembl 96) using STAR (version 2.5.3a)^59^. The number of mapped reads were counted for each gene and compiled into an expression matrix using featureCounts (version 1.6.0)^60^. This resulted in a table with gene counts for 58,929 genes. Further analysis was performed in Rstudio (version 2023.03.1+446)^61^. As an additional quality control step, variance stabilising transformation (vst)-normalised data was analysed using principal component analysis to identify potential outliers; there was no evidence of outliers.

Experimental conditions were analysed for differentially expressed genes (DEGs) using the DESeq2 package (version 1.40.2)^62^ and the lfcShrink apeglm method to obtain log2FC for further analysis^63^. For TMAO and TMA, a DEG cut-off of adjusted p-value<0.05 was used. For MeHg and NaAs, a log2FC ≥0.5 was added as an additional criterion due to the large number of DEGs.

DAVID (consulted on 19 July 2024, version 2021)^64,65^ was used to perform Gene Ontology (GO) analysis (Biological processes, GO FAT) against a background of genes with at least one count in any of the exposure conditions (42816 genes of which 27418 recognised by DAVID). Synaptic GO-term enrichment was explored using the SynGO platform (version 1.2)^66^ with the same set of background genes as for DAVID. To determine whether Kyoto Encyclopedia of Genes and Genomes (KEGG) pathways were activated or inhibited in exposed cells, Signaling Pathway Impact Analysis (SPIA) was performed on the DEGs^67^. For this analysis DEGs of MeHg with an adjusted p-value of <0.05 were taken to consider subtle changes as well, for NaAs the FC cut-off of 0.5 was kept due to the large numbers of DEGs. Human KEGG pathways were considered significantly regulated when p_GFDR_ was ≤0.05 for all compounds except for TMA (≤0.3). The pathways were downloaded in KMGL format from the KEGG PATHWAY database (download date: 19 Oct 2022) and were used to do the network analysis (KEGGgraph, RBGL^68^). Gene interactions were analysed in STRING (version 12.0, full network, medium confidence (0.4))^69^ and colour coded according to the role of those genes in specific pathways.

Gene expression heatmaps (hierarchical clustering Euclidean distance, ward.D linkage) and PCA plots were made in RStudio, the Venn diagram was produced using Venny (version 2.1.0)^70^, ShinyGO was used to visualise enriched GO-terms^71^, SynGO plots were made in the custom SynGO plot tool, and STRING interactions were visualised in Cytoscape (version 3.7.4)^72^, using perfuse force-directed layout. Other graphs were visualised using GraphPad Prism (version 10.5.0).

#### Multi-well microelectrode array (MEA) measurements

Spontaneous electrical activity was measured as described before^54^. Briefly, spontaneous electrical activity was measured every three to four days starting from three days after the initiation of neuronal differentiation. Cultures were grown in PEI-coted 48-well MEA plates, each well containing 16 nanotextured gold microelectrodes, yielding a total of 768 channels (Axion Biosystems Inc., Atlanta, GA, USA). For every measurement, the mwMEA plate was placed into the MaestroPro (incubator set at 37°C, 5% CO_2_) for at least 15 min before starting a recording of spontaneous activity of 15 min. Data was acquired using the Axion Integrated Studio (AxIS) Navigator (version 3.10.3.). Channels were sampled at the default rate of 12.5 kHz and the resulting signal were band-pass filtered at 0.2–3 kHz. Raw data was re-recorded and spikes detected with the AxIS Spike Detector (Adaptive threshold crossing) using a spike duration of 0.84/2.16 ms (pre/post) and a spike threshold of 5.5 SD above the internal noise level. An electrode was considered active when it fired at least 5 spikes/min, bursts were detected with the Poisson surprise method (> 10 surprises). Network bursts were detected using the adaptive setting, with the minimum number of spikes set at 50 and minimum percentage of electrodes at 15%. Data was exported using the Axion Neural Metrics Tool (version 4.1.1) and Axion Metric Plotting Tool (version 2.5.1). Datasets were obtained from two independent experiments with eight technical replicates per experimental condition, in line with published guidelines^24^. All metrics were normalised by dividing each timepoint by the average over all timepoints. Statistical analysis and visualisation was done using GraphPad Prism (version 10.5.0), using a two-way ANOVA and Tukey’s multiple comparisons test, and the Area Under the Curve calculation tool.

## Acknowledgements

This project has received funding from the European Union’s Horizon 2020 research and innovation programme under grant agreement No. 874583 (ATHLETE project). This publication reflects only the authors’ views and the European Commission is not responsible for any use that may be made of the information it contains. We are grateful to all the participating families, practitioners, schools and researchers in France, Greece, Lithuania, Spain, Norway and the UK who made this study happen. This project is only possible because of their enthusiasm and commitment.

VL, CO and EH were additionally funded by the Dutch Ministry for Public Health and Health Care programme 5.1.2. We would like to thank Yvonne Staal and Cristina Villanueva for a critical review of the manuscript. We acknowledge support from the grant CEX2023-0001290-S funded by MCIN/AEI/ 10.13039/501100011033, and support from the Generalitat de Catalunya through the CERCA Program and from AGAUR SGR 2021_PI4970_2021 SGR 01576.

LM has received funding from the Ramon y Cajal Research Fellowship RYC2022-036475-I funded by MICIU/AEI/10.13039/501100011033. INMA data collections were supported by grants from the Instituto de Salud Carlos III, CIBERESP, and the Generalitat de Catalunya-CIRIT.

## Resource availability

### Lead contact

Requests for further information and resources should be directed to and will be fulfilled by the lead contact, Simon McArthur.

### Materials availability

This study did not generate new unique reagents.

### Data and code availability

Data:

– HELIX and INMA data upon request
– RNAseq data: Gene expression data have been deposited in the NCBI GEO database under accession number E-MTAB-16884 (TMAO, TMA, NaAs) and GSE166297 (MeHg).
– MEA data – https://doi.org/10.6084/m9.figshare.31076257

Code:

– All original analytical code for the epidemiology analyses has been deposited in a publicly available Github repository. https://github.com/HERMES-Exposome/TMAO_exposure_neurodevelopment.git

## Authors’ contributions

Conceptualisation was the responsibility of LM, JJ, HK, EH, LH, SMc

Coordination was the responsibility of HK, MV

Methodology and investigation were the responsibility of CO, LM, VL

Formal analysis was conducted by VL, LM, ERD

Writing was carried out by VL, LM, LH, ERD, EH, LH, SMc

Review of the manuscript was done by VL, LM, CO, ERD, AA, LC, MC, RG, BH, JI, JJ, HK, AP, LM, SM, MRR, MS-P, AB, MT, MV, JW, EH, LH, SMc

Supervision of the study was carried out by LH, SMc, EH

Funding acquisition was the responsibility of HK, LH, EH, MV

## References

1. MetaHIT Consortium et al. A human gut microbial gene catalogue established by metagenomic sequencing. Nature 464, 59–65 (2010).

2. Caspani, G. & Swann, J. Small talk: microbial metabolites involved in the signaling from microbiota to brain. Curr. Opin. Pharmacol. 48, 99–106 (2019).

3. Leader, G. et al. Gastrointestinal Symptoms in Autism Spectrum Disorder: A Systematic Review. Nutrients 14, 1471 (2022).

4. Wan, Y. et al. Underdevelopment of the gut microbiota and bacteria species as non-invasive markers of prediction in children with autism spectrum disorder. Gut 71, 910–918 (2022).

5. Lou, M. et al. Deviated and early unsustainable stunted development of gut microbiota in children with autism spectrum disorder. Gut 71, 1588–1599 (2022).

6. Zuffa, S. et al. Early-life differences in the gut microbiota composition and functionality of infants at elevated likelihood of developing autism spectrum disorder. Transl. Psychiatry 13, 257 (2023).

7. Morton, J. T. et al. Multi-level analysis of the gut–brain axis shows autism spectrum disorder-associated molecular and microbial profiles. Nat. Neurosci. 26, 1208–1217 (2023).

8. Silva, Y. P., Bernardi, A. & Frozza, R. L. The Role of Short-Chain Fatty Acids From Gut Microbiota in Gut-Brain Communication. Front. Endocrinol. 11, 25 (2020).

9. Gao, K., Mu, C., Farzi, A. & Zhu, W. Tryptophan Metabolism: A Link Between the Gut Microbiota and Brain. Adv. Nutr. 11, 709–723 (2020).

10. Hoyles, L. et al. Regulation of blood–brain barrier integrity by microbiome-associated methylamines and cognition by trimethylamine N-oxide. Microbiome 9, 235 (2021).

11. Jimenez-Arenas, P. et al. The relationship between the gut microbiota and neuropsychological development and behaviour during childhood and adolescence: a systematic review of epidemiological studies. Brain Behav. Immun. – Health 53, 101212 (2026).

12. Yap, C. X. et al. Autism-related dietary preferences mediate autism-gut microbiome associations. Cell 187, 495–510 (2024).

13. Wu, Y. et al. The Mediating Role of Eating Behaviors Between Autistic Symptoms and Dietary Issues Among Chinese Children With Autism. J. Autism Dev. Disord. https://doi.org/10.1007/s10803-025-07133-y (2025) doi:10.1007/s10803-025-07133-y.

14. Zhang, S., Han, F., Wang, Q. & Fan, F. Probiotics and Prebiotics in the Treatment of Autism Spectrum Disorder: A Narrative Review. J. Integr. Neurosci. 23, 20 (2024).

15. Sandler, R. H. et al. Short-Term Benefit From Oral Vancomycin Treatment of Regressive-Onset Autism. J. Child Neurol. 15, 429–435 (2000).

16. Maitre, L. et al. Multi-omics signatures of the human early life exposome. Nat. Commun. 13, 7024 (2022).

17. Maitre, L. et al. Urine Metabolic Signatures of Multiple Environmental Pollutants in Pregnant Women: An Exposome Approach. Environ. Sci. Technol. 52, 13469–13480 (2018).

18. Krüger, R. et al. Associations of current diet with plasma and urine TMAO in the KarMeN study: direct and indirect contributions. Mol. Nutr. Food Res. 61, 1700363 (2017).

19. Turunen, A. W. et al. Dioxins, polychlorinated biphenyls, methyl mercury and omega-3 polyunsaturated fatty acids as biomarkers of fish consumption. Eur. J. Clin. Nutr. 64, 313–323 (2010).

20. Vuong, H. E. et al. The maternal microbiome modulates fetal neurodevelopment in mice. Nature 586, 281–286 (2020).

21. De Leeuw, V. C. et al. Neuronal differentiation pathways and compound-induced developmental neurotoxicity in the human neural progenitor cell test (hNPT) revealed by RNA-seq. Chemosphere 304, 135298 (2022).

22. Human Metabolite Database. https://hmdb.ca/metabolites/HMDB000092; https://hmdb.ca/metabolites/HMDB0000906. Accessed on 25 May 2025

23. European Human Biomonitoring Dashboard. https://hbm.vito.be/eu-hbm-dashboard. Accessed on 17 Nov 2025

24. Mossink, B. et al. Human neuronal networks on micro-electrode arrays are a highly robust tool to study disease-specific genotype-phenotype correlations in vitro. Stem Cell Rep. 16, 2182–2196 (2021).

25. Bragg, M. G. et al. The association between plasma choline, growth and neurodevelopment among Malawian children aged 6–15 months enroled in an egg intervention trial. Matern. Child. Nutr. 19, e13471 (2023).

26. Padilha, M. et al. Serum metabolome indicators of early childhood development in the Brazilian National Survey on Child Nutrition (ENANI-2019). eLife 14, e97982 (2025).

27. Chilloux, J. et al. Inhibition of IRAK4 by microbial trimethylamine blunts metabolic inflammation and ameliorates glycemic control. Nat. Metab. 7, 2531–2547 (2025).

28. Finegan, J.-A., Bartleman, B. & Wong, P. Y. A Window for the Study of Prenatal Sex Hormone Influences on Postnatal Development. J. Genet. Psychol. 150, 101–112 (1989).

29. Savic, I., Garcia-Falgueras, A. & Swaab, D. F. Sexual differentiation of the human brain in relation to gender identity and sexual orientation. in Progress in Brain Research vol. 186 41–62 (Elsevier, 2010).

30. Ripp, S. L., Itagaki, K., Philpot, R. M. & Elfarra, A. A. Species and Sex Differences in Expression of Flavin-Containing Monooxygenase Form 3 in Liver and Kidney Microsomes. Drug Metab. Dispos. 27, 46–52 (1999).

31. Hines, R. N. Developmental expression of drug metabolizing enzymes: Impact on disposition in neonates and young children. Int. J. Pharm. 452, 3–7 (2013).

32. Aaseth, J., Wallace, D. R., Vejrup, K. & Alexander, J. Methylmercury and developmental neurotoxicity: A global concern. Curr. Opin. Toxicol. 19, 80–87 (2020).

33. Tolins, M., Ruchirawat, M. & Landrigan, P. The Developmental Neurotoxicity of Arsenic: Cognitive and Behavioral Consequences of Early Life Exposure. Ann. Glob. Health 80, 303 (2014).

34. Wang, S. et al. Trimethylamine-N-oxide disrupts spermatogenesis by inducing mitochondrial oxidative stress injury through Hippo signaling. Free Radic. Biol. Med. 243, 452–465 (2026).

35. Bordoni, L., Petracci, I. & Gabbianelli, R. TMA, beyond TMAO, might contribute to vascular inflammation by disturbing mitochondrial functions in macrophages. Biochem. Biophys. Res. Commun. 754, 151529 (2025).

36. Wang, J., Gu, X., Yang, J., Wei, Y. & Zhao, Y. Gut Microbiota Dysbiosis and Increased Plasma LPS and TMAO Levels in Patients With Preeclampsia. Front. Cell. Infect. Microbiol. 9, 409 (2019).

37. Wen, Y. et al. Maternal serum trimethylamine-N-oxide is significantly increased in cases with established preeclampsia. Pregnancy Hypertens. 15, 114–117 (2019).

38. Huang, X. et al. Association between risk of preeclampsia and maternal plasma trimethylamine-N-oxide in second trimester and at the time of delivery. BMC Pregnancy Childbirth 20, 302 (2020).

39. Jääskeläinen, T., Kärkkäinen, O., Heinonen, S., Hanhineva, K. & Laivuori, H. No association in maternal serum levels of TMAO and its precursors in pre-eclampsia and in non-complicated pregnancies. Pregnancy Hypertens. 28, 74–80 (2022).

40. Andrikopoulos, P. et al. Evidence of a causal and modifiable relationship between kidney function and circulating trimethylamine N-oxide. Nat. Commun. 14, 5843 (2023).

41. Li, Q. et al. Synchronous Evolution of an Odor Biosynthesis Pathway and Behavioral Response. Curr. Biol. 23, 11–20 (2013).

42. Haug, L. S. et al. In-utero and childhood chemical exposome in six European mother-child cohorts. Environ. Int. 121, 751–763 (2018).

43. Maitre, L. et al. Early-life environmental exposure determinants of child behavior in Europe: A longitudinal, population-based study. Environ. Int. 153, 106523 (2021).

44. Tamayo-Uria, I. et al. The early-life exposome: Description and patterns in six European countries. Environ. Int. 123, 189–200 (2019).

45. Vrijheid, M. et al. The Human Early-Life Exposome (HELIX): Project Rationale and Design. Environ. Health Perspect. 122, 535–544 (2014).

46. Achenbach, T. M. & Rescorla, L. A. Child Behavior Checklist for Ages 6-18. 10.1037/t47452-000 (2023).

47. Conners, C. M. Conners’ Rating Scales — Revised: User’s Manual. (Multi-Health Systems, North Tonawanda, New York, 1997).

48. Raven, J., Raven, J. & Court, J. Section 4: The advanced progressive matrices. in Manual for Raven’s progressive matrices and vocabulary scales (Oxford Psychologists Press., Oxford, 1998).

49. Forns, J. et al. The n-back Test and the Attentional Network Task as measures of child neuropsychological development in epidemiological studies. Neuropsychology 28, 519–529 (2014).

50. Julvez, J. et al. Early life multiple exposures and child cognitive function: A multi-centric birth cohort study in six European countries. Environ. Pollut. 284, 117404 (2021).

51. Maitre, L. et al. Assessment of metabolic phenotypic variability in children’s urine using 1H NMR spectroscopy. Sci. Rep. 7, 46082 (2017).

52. Lau, C.-H. E. et al. Determinants of the urinary and serum metabolome in children from six European populations. BMC Med. 16, 202 (2018).

53. Calvo-Serra, B. et al. Urinary metabolite quantitative trait loci in children and their interaction with dietary factors. Hum. Mol. Genet. 29, 3830–3844 (2021).

54. De Leeuw, V. C. et al. An efficient neuron-astrocyte differentiation protocol from human embryonic stem cell-derived neural progenitors to assess chemical-induced developmental neurotoxicity. Reprod. Toxicol. 98, 107–116 (2020).

55. Gunhanlar, N. et al. A simplified protocol for differentiation of electrophysiologically mature neuronal networks from human induced pluripotent stem cells. Mol. Psychiatry 23, 1336–1344 (2018).

56. Slob, W. Dose-Response Modeling of Continuous Endpoints. Toxicol. Sci. 66, 298–312 (2002).

57. R Core Team. R: A Language and Environment for Statistical Computing. R Foundation for Statistical Computing.

58. Ewels, P., Magnusson, M., Lundin, S. & Käller, M. MultiQC: summarize analysis results for multiple tools and samples in a single report. Bioinformatics 32, 3047–3048 (2016).

59. Dobin, A. et al. STAR: ultrafast universal RNA-seq aligner. Bioinformatics 29, 15–21 (2013).

60. Liao, Y., Smyth, G. K. & Shi, W. featureCounts: an efficient general purpose program for assigning sequence reads to genomic features. Bioinformatics 30, 923–930 (2014).

61. Posit team. RStudio: Integrated Development Environment for R.

62. Love, M. I., Huber, W. & Anders, S. Moderated estimation of fold change and dispersion for RNA-seq data with DESeq2. Genome Biol. 15, 550 (2014).

63. Zhu, A., Ibrahim, J. G. & Love, M. I. Heavy-tailed prior distributions for sequence count data: removing the noise and preserving large differences. Bioinformatics 35, 2084–2092 (2019).

64. Huang, D. W., Sherman, B. T. & Lempicki, R. A. Systematic and integrative analysis of large gene lists using DAVID bioinformatics resources. Nat. Protoc. 4, 44–57 (2009).

65. Sherman, B. T. et al. DAVID: a web server for functional enrichment analysis and functional annotation of gene lists (2021 update). Nucleic Acids Res. 50, W216–W221 (2022).

66. Koopmans, F. et al. SynGO: An Evidence-Based, Expert-Curated Knowledge Base for the Synapse. Neuron 103, 217–234.e4 (2019).

67. Tarca, A. L. et al. A novel signaling pathway impact analysis. Bioinformatics 25, 75–82 (2009).

68. Zhang, J. D. & Wiemann, S. KEGGgraph: a graph approach to KEGG PATHWAY in R and bioconductor. Bioinformatics 25, 1470–1471 (2009).

69. Szklarczyk, D. et al. The STRING database in 2023: protein–protein association networks and functional enrichment analyses for any sequenced genome of interest. Nucleic Acids Res. 51, D638–D646 (2023).

70. Oliveros, J. Venny. An interactive tool for comparing lists with Venn’s diagrams.

71. Ge, S. X., Jung, D. & Yao, R. ShinyGO: a graphical gene-set enrichment tool for animals and plants. Bioinformatics 36, 2628–2629 (2020).

72. Shannon, P. et al. Cytoscape: A Software Environment for Integrated Models of Biomolecular Interaction Networks. Genome Res. 13, 2498–2504 (2003).

